# Genetically proxied inhibition of L-2-hydroxyglutarate dehydrogenase and the risk of coronary artery disease: A Mendelian randomization study

**DOI:** 10.1101/2023.01.15.23284550

**Authors:** Euijun Song

## Abstract

**Background:** L-2-hydroxyglutarate dehydrogenase (L2HGDH) deletion-induced L-2-hydroxyglutarate accumulation plays a cardioprotective role in hypoxic conditions. However, there has been no causal evidence in real-world clinical data. We aimed to examine the causal effects of *L2HGDH* inhibition on coronary artery disease (CAD) and myocardial infarction (MI) using Mendelian randomization (MR) analysis.

**Methods:** We used nine *L2HGDH*-proxied genetic variants associated with blood 2-hydroxyglutarate levels as genetic instruments, and performed two-sample MR analysis using the CARDIoGRAMplusC4D meta-analysis datasets of CAD (60,801 CAD cases and 123,504 controls) and MI (34,541 MI cases and 261,984 controls).

**Results:** Genetically proxied inhibition of *L2HGDH* associated with 2-hydroxyglutarate levels potentially decreased the risk of CAD (odds ratio [OR] 0.486, 95% confidence interval [CI] 0.242–0.977, P=0.043) but was not associated with the risk of MI (OR 0.676, 95% CI 0.312–1.463, P=0.320). This potentially causal association between *L2HGDH* inhibition and CAD was unlikely to be biased by horizontal pleiotropy, whereas there might be a weak instrument bias.

**Conclusion:** Our MR analysis suggests the potential association between genetically proxied inhibition of *L2HGDH* and CAD. Our findings may have therapeutic implications for L2HGDH inhibitors in CAD, and further large-scale clinical studies are needed.

## 1. Introduction

L-2-hydroxyglutarate plays a pivotal role in cytoplasmic and mitochondrial energy metabolism to maintain redox regulation [1]. Hypoxia increases cellular L-2-hydroxyglutarate, which is produced from α-ketoglutarate by malate dehydrogenase [1-3]. L-2-hydroxyglutarate dehydrogenase (L2HGDH) is the only enzyme known to oxidize L-2-hydroxyglutarate back to α-ketoglutarate [2, 4]. In experiments, *L2HGDH* ablation increases the accumulation of L-2-hydroxyglutarate in hypoxia [2, 5]. L-2-hydroxyglutarate has been known to protect heart tissue from oxidative stress in response to hypoxia [5]. Since coronary artery disease (CAD) leads to ischemic injury and ischemia-reperfusion injury, L-2-hydroxyglutarate may play a protective role in CAD and ischemic myocardial injury.

Metabolic remodeling plays a key role in the pathobiology of CAD and myocardial infarction (MI) [5-7]. Recently, He and colleagues [5] discovered that genetic *L2HGDH* deletion-induced L-2-hydroxyglutarate accumulation shows cardioprotective effects under ischemic conditions in mice. They found that L-2-hydroxyglutarate accumulation shifts glucose metabolisms from glycolysis to the pentose phosphate pathway in response to low-flow ischemia or ischemia-reperfusion. This metabolic shift indicates that *L2HGDH* deletion-induced L-2-hydroxyglutarate accumulation could reduce ischemic myocardial damage by eliminating reactive oxygen species. The experiments suggest that *L2HGDH* is a potential therapeutic target for oxidative stress-related cardiovascular diseases, such as CAD and MI. However, there has been no prospective clinical trial yet.

One genetic epidemiological approach for testing the causal effects of drug targets on complex diseases is the Mendelian randomization (MR) method [8-12]. MR methods can estimate the causal effects of putative risk factors based on germline genetic variation in the population, without conducting a randomized controlled trial. Using the drug-target MR framework, Ference and colleagues [8] determined the causal associations of ATP citrate lyase (ACLY) and HMG-CoA reductase (HMGCR) inhibitions with the risk of cardiovascular disease. Similarly, MR approaches could identify the potentially causal associations of genetically proxied inhibition of cardiovascular drug targets with cancers, neuropsychiatric diseases, and cognitive function [9-12].

Here, we conduct a two-sample MR study to assess putative causal effects of genetically proxied inhibition of *L2HGDH* on CAD and MI (Figure 1). We construct genetic instruments for *L2HGDH* that are associated with blood 2-hydroxyglutarate levels. We then perform MR analysis to test whether this genetic instrument is causally associated with CAD and MI using large-scale genome-wide association study (GWAS) datasets.

**Figure 1.**
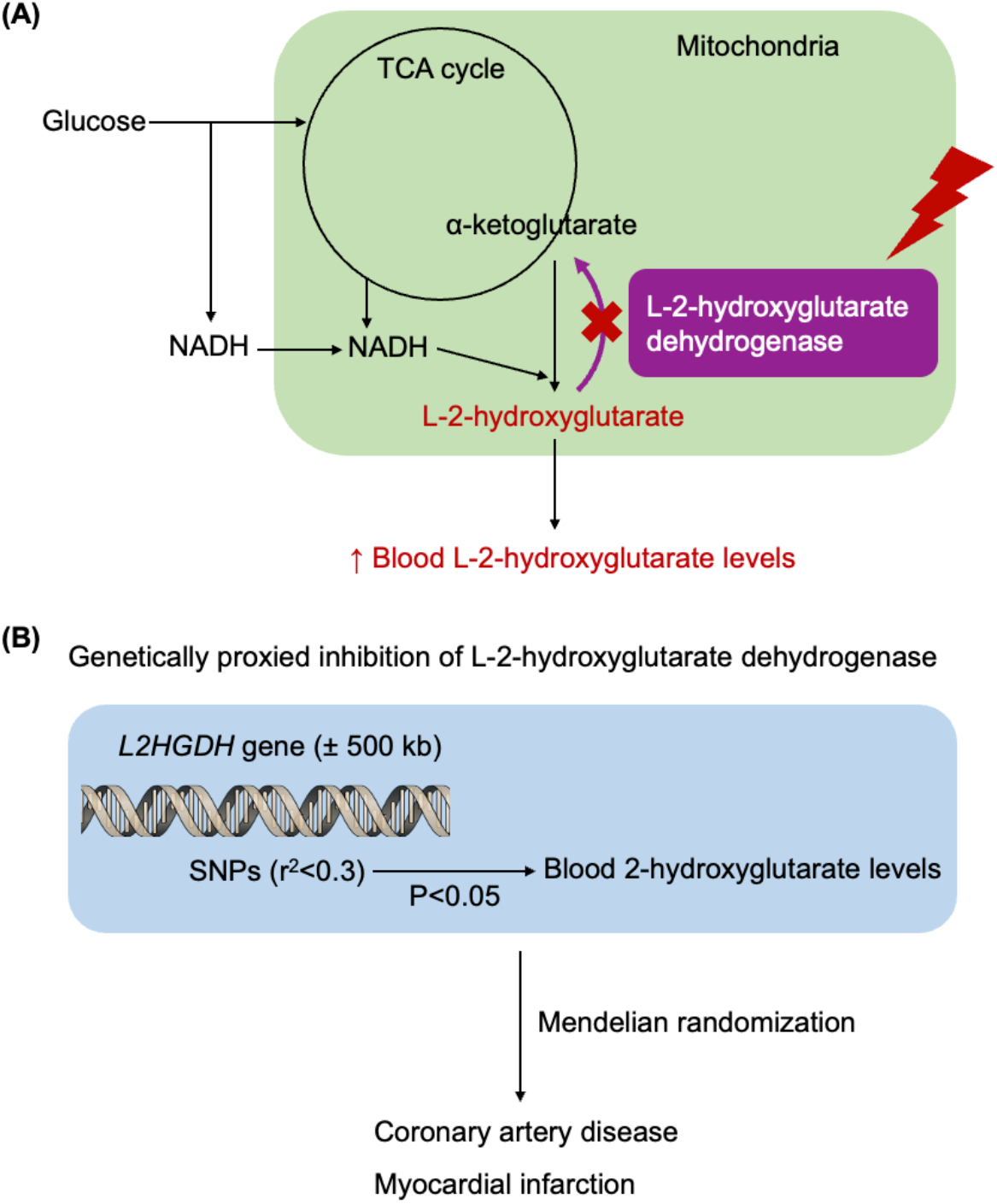
Overall analysis scheme. (A) Function of L-2-hydroxyglutarate dehydrogenase (L2HGDH) in energy metabolisms. Inhibition of L2HGDH increases blood L-2-hydroxyglutarate levels. (B) We constructed genetic instruments proxied to the *L2HGDH* gene that are associated with blood 2-hydroxyglutarate levels. We then performed two-sample Mendelian randomization analysis to assess the causal effects of genetically proxied inhibition of *L2HGDH* on coronary artery disease and myocardial infarction.

## 2. Methods

### 2.1. Mendelian randomization study design

As illustrated in Figure 1, we designed a two-sample MR study to determine putative causal effects of genetically proxied inhibition of *L2HGDH* on CAD and MI. We first constructed the genetic instruments for *L2HGDH* to mimic pharmacological/genetic perturbation of *L2HGDH* for increasing L-2-hydroxyglutarate levels. Due to the limited availability of GWAS data for L-2-hydroxyglutarate levels, we constructed the genetic instruments that are associated with 2-hydroxyglutarate level instead. We then performed the Mendelian randomization analysis to determine potential causal associations between the genetic instruments for *L2HGDH* and CAD using large-scale GWAS summary statistics datasets.

### 2.2. Genetic instruments for genetically proxied inhibition of *L2HGDH*

We obtained the GWAS summary statistics for human blood metabolome from Shin et al [13]. Shin et al. [13] analyzed >400 blood metabolites from the KORA and TwinsUK European population datasets using the Metabolon platform. Since the Metabolon platform could not distinguish L-2-hydroxyglutarate from D-2-hydroxyglutarate, we alternatively used the GWAS data for 2-hydroxyglutarate levels (accessed from https://gwas.mrcieu.ac.uk/) to construct the genetic instruments for *L2HGDH*.

L2HGDH is the only enzyme known to catabolize L-2-hydroxyglutarate in human cells, and L2HGDH inhibition increases L-2-hydroxyglutarate levels in experiments [1, 2]. Since 2-hydroxyglutarate includes both L-2-hydroxyglutarate and D-2-hydroxyglutarate, we only considered *L2HGDH*-proxied genetic variants to mainly reflect L-2-hydroxyglutarate levels. We constructed the genetic instruments for *L2HGDH* by recruiting genetic variants within ± 500 kb from the *L2HGDH* gene that are associated with blood 2-hydroxyglutarate level (P<0.05) and that are also in low linkage disequilibrium (r^2^<0.3, with the default clumping window size of 10000 kb) [8]. The characteristics of genetic variants proxied to *L2HGDH* are shown in Table S1. Since there was no genome-wide significant variant near the *L2HGDH* gene (Table S2), we adopted weak P-value and r^2^ thresholds previously suggested by Ference and colleagues [8]. The SNPs and *L2HGDH* gene were analyzed based on the GRCh37 (hg19) human genome reference (Figure S1). The PhenoScanner V2 (http://www.phenoscanner.medschl.cam.ac.uk/, accessed on Feb 6, 2023) was used to identify pleiotropic variants (Table S3).

### 2.3. Data sources for CAD and MI

The GWAS summary statistics for CAD was obtained from the CARDIoGRAMplusC4D (Coronary Artery DIsease Genome-wide Replication and Meta-analysis plus Coronary Artery Disease) 1000 Genomes-based GWAS meta-analysis study (http://www.cardiogramplusc4d.org) [14]. The CARDIoGRAMplusC4D GWAS meta-analysis dataset included genetic associations from 60,801 CAD cases and 123,504 controls mostly of European ancestry. The case status was defined by diagnosis of CAD, including MI, acute coronary syndrome, chronic stable angina, and coronary stenosis >50% [14]. In addition, the MI GWAS summary statistics was obtained from the same CARDIoGRAMplusC4D meta-analysis study (34,541 MI cases and 261,984 controls). All the MI cases are a subpopulation of the CAD cases [14].

### 2.4. Mendelian randomization and statistical analysis

For MR analysis, the instrument strength was measured using the F-statistic. The F-statistic less than 10 indicates weak instruments. We used the inverse-variance weighted (IVW), weighted median, maximum likelihood, MR-PRESSO [15], Radial-MR [16], and MR-RAPS [17] methods to evaluate the putative causal effects of genetically proxied inhibition of *L2HGDH* on CAD and MI. The heterogeneity between SNPs was evaluated by Cochran’s Q test [18]. We also performed the MR Steiger directionality test to determine the directionality of causality [19]. To assess potential directional and horizontal pleiotropy, we used the MR-Egger intercept test and MR-PRESSO methods. The leave-one-out sensitivity analysis and Radial-MR were used to detect potential outlying genetic variants. P<0.05 was considered statistically significant. All MR analyses were performed in R (version 4.1.2, https://www.r-project.org/) with the packages TwoSampleMR, MRPRESSO, and RadialMR.

## 3. Results

### 3.1. Genetic instruments for *L2HGDH*

We first constructed the genetic instruments proxied to *L2HGDH* that are associated with blood 2-hydroxyglutarate levels (P<0.05) and that are also in low linkage disequilibrium (r^2^<0.3; Table S1). Since there was no genome-wide significant variant associated with 2-hydroxyglutarate levels near the *L2HGDH* gene (Table S2), we adopted a loose significance threshold of 0.05 to construct instruments for genetically proxied inhibition of *L2HGDH* [8]. These genetic instruments mimic genetic or pharmacological inhibition of *L2HGDH* for increasing blood 2-hydroxyglutarate levels (Figure 1). The genetic instruments have the mean F-statistic of 7.661, indicating that there might be a weak instrument bias in our MR analysis. The genetic instruments are not associated with possible confounders or cardiovascular diseases/traits (Table S3).

### 3.2. Association of genetically proxied inhibition of *L2HGDH* with CAD and MI

We performed a two-sample MR analysis using the CARDIoGRAMplusC4D GWAS meta-analysis dataset of CAD (60,801 CAD cases and 123,504 controls). As highlighted in Table 1, the IVW method showed that genetically proxied inhibition of *L2HGDH* associated with 2-hydroxyglutarate levels potentially decreased the risk of CAD (OR 0.486, 95% CI 0.242–0.977, P=0.043). There was no significant evidence of heterogeneity (Cochran’s Q-statistic=5.757, P=0.674). The radial IVW, weighted median, and maximum likelihood methods also showed directionally similar estimates (Table 1). The MR-RAPS method indicated that there might be a slightly weak instrument bias (OR 0.460, 95% CI 0.202–1.046, P=0.064). The MR Steiger directionality test confirmed the direction of the causality (P<0.001).

**Table 1.** Mendelian randomization (MR) analyses for causal associations of genetically proxied inhibition of *L2HGDH* with coronary artery disease and myocardial infarction.

|  | **Coronary artery disease**<br>(60,801 cases, 123,504 controls) |  |  | **Myocardial infarction**<br>(34,541 cases, 261,984 controls) |  |  |
| --- | --- | --- | --- | --- | --- | --- |
|  | **OR (95% CI)** | **P-value** | **SNPs** | **OR (95% CI)** | **P-value** | **SNPs** |
| IVW | 0.486 (0.242–0.977) | 0.043 | 9 | 0.676 (0.312–1.463) | 0.320 | 9 |
| IVW (Radial) | 0.487 (0.269–0.880) | 0.017 | 9 | 0.676 (0.361–1.268) | 0.222 | 9 |
| Weighted median | 0.762 (0.300–1.937) | 0.568 | 9 | 0.888 (0.325–2.428) | 0.817 | 9 |
| Maximum likelihood | 0.457 (0.216–0.967) | 0.041 | 9 | 0.655 (0.291–1.476) | 0.308 | 9 |
| MR-RAPS | 0.460 (0.202–1.046) | 0.064 | 9 | 0.681 (0.284–1.630) | 0.388 | 9 |
| MR-Egger intercept* | -0.014 (0.016) | 0.396 | 9 | -0.004 (0.017) | 0.836 | 9 |
| MR-PRESSO global<sup>#</sup> | - | 0.715 | 9 | - | 0.752 | 9 |
| MR Steiger directionality | - | <0.001 | 9 | - | <0.001 | 9 |

SNP, single nucleotide polymorphism; OR, odds ratio; CI, confidence interval; IVW, inverse-variance weighted method.
\*Intercept values are presented with SE.
<sup>#</sup>No outlying genetic variants were detected.

To assess potential directional horizontal pleiotropy, we used the MR-Egger and MR-PRESSO methods. The MR-Egger intercept test determined that 9 genetic variants for *L2HGDH* showed no evidence for significant pleiotropy (intercept=-0.014, P=0.396). The MR-PRESSO global test also confirmed no significant horizontal pleiotropy with no outlying genetic variants (P=0.715). In addition, the leave-one-out sensitivity analysis showed that removing individual genetic variants did not substantially change the estimated causal effect of genetically proxied inhibition of *L2HGDH* (Table S4). The radial IVW method further confirmed no evidence of outlying genetic variants (Figure S2). Taken together, the sensitivity analyses provide no significant evidence for pleiotropic effects, which could bias the MR analysis.

As a subpopulation of the CAD cases, we also examined the putative causal association between *L2HGDH* inhibition and MI using the CARDIoGRAMplusC4D meta-analysis dataset of MI (34,541 MI cases and 261,984 controls). Using IVW, we found that genetically proxied inhibition of *L2HGDH* was not significantly associated with the risk of MI (OR 0.676, 95% CI 0.312–1.463, P=0.320). The MR-Egger intercept and MR-PRESSO global tests indicated no significant pleiotropy (Table 1), and the radial plot indicated no evidence of outlying variants (Figure S2).

## 4. Discussion

To our knowledge, this is the first MR study to demonstrate the potentially causal association between genetically proxied inhibition of *L2HGDH* and CAD. Using two-sample MR analyses, we revealed that genetically proxied inhibition of *L2HGDH* was significantly associated with the decreased risk of CAD. The multiple MR sensitivity analyses confirmed that this potential causal association was unlikely to be biased by horizontal pleiotropy. Our findings suggests that genetic/pharmacological targeting of *L2HGDH* may decrease the risk of CAD by increasing blood 2-hydroxyglutarate levels. Since our genetic instruments are not directly associated with L-2-hydroxyglutarate levels and there might be a weak instrument bias, further large-scale GWAS studies are needed to obtain more robust and reliable MR results.

In the previous experimental study, *L2HGDH* deletion-induced L-2-hydroxyglutarate accumulation protects the heart from redox stress, which plays a key role in myocardial ischemic injury [5]. However, there has been no clinical trial to validate this potential cardioprotective effect of *L2HGDH* inhibition. Using large-scale MR, we revealed that genetically proxied inhibition of *L2HGDH* could potentially decrease the risk of CAD but was not significantly associated with MI *per se* (Table 1). In our MR study, the CAD cases include MI, acute coronary syndrome, coronary atherosclerosis, and chronic ischemic heart diseases. Therefore, our findings suggest that *L2HGDH* inhibition may play a cardioprotective role in not only being limited to myocardial ischemic damage, but also in the primary prevention of CAD. The mechanisms underlying the potential preventive effects of *L2HGDH* inhibition on CAD should be further investigated.

Due to the limited availability of GWAS data on blood metabolome, we constructed genetic instruments based on blood 2-hydroxyglutarate levels, including its two enantiomers L-2-hydroxyglutarate and D-2-hydroxyglutarate. D-2-hydroxyglutarate has been known to be an oncometabolite in certain types of rare cancers [20, 21]. To minimize the effects of D-2-hydroxyglutarate in our MR analysis, we constructed the genetic instruments only proxied to the *L2HGDH* gene. In experiments, L-2-hydroxyglutarate is the majority of 2-hydroxyglutarate, and *L2HGDH* but not D2HGDH knockdown increases the concentration of 2-hydroxyglutarate [1, 2]. Additionally, L2HGDH is the only reported enzyme that metabolizes L-2-hydroxyglutarate. Therefore, our approach would mainly reflect *L2HGDH* inhibition-related L-2-hydroxyglutarate levels; however, large-scale GWAS data of blood L-2-hydroxyglutarate levels is essential to obtain more robust and reliable MR results. Since L-2-hydroxyglutarate is also a putative oncometabolite in renal cell carcinoma [22, 23], further epidemiological/clinical studies are warranted to determine the potential side effects of *L2HGDH* inhibition.

A critical limitation of this study is that we adopted a loose P-value threshold of 0.05 to build the genetic instruments [8]. When we used the genome-wide significance threshold, there was no genetic variant associated with blood 2-hydroxyglutarate levels, near the *L2HGDH* gene (Table S2). Since L2HGDH is the only enzyme known to metabolize L-2-hydroxyglutarate, we only considered *L2HGDH*-proxied variants to reflect the physiological effects of *L2HGDH* inhibition on L-2-hydroxyglutarate levels [1, 2]. In addition, we did not perform any replication analysis in non-European populations, though the CARDIoGRAMplusC4D is the largest consortium study of CAD. Another critical limitation is the underlying overlap between exposure and outcome datasets, as the CARDIoGRAMplusC4D includes KORA (also known as GerMIFS III) and TwinsUK (part of the PROCARDIS) datasets. This overlap could cause a bias towards confounded associations in the two-sample MR [24]. In our MR analysis, the weighted median methods did not show significant results; however, those methods are statistically less powerful than IVW [25]. Large-scale multi-ancestry GWAS datasets for L-2-hydroxyglutarate are essential to construct more robust genetic instruments. Protein-protein interaction network analysis would be helpful to determine the functional relationship between L2HGDH and the CAD disease module and to identify potential drug targets [26, 27].

## 5. Conclusion

Our MR result suggests the potential association between genetically proxied inhibition of *L2HGDH* and CAD, though there might be a weak instrument bias. Our findings may have therapeutic implications for L2HGDH inhibitors in CAD. Further large-scale clinical studies are needed to obtain more robust results.

## Data Availability

All data produced in the present study are available upon reasonable request to the authors.

## Authorship contribution

Euijun Song designed, performed, analyzed data, and wrote the manuscript.

## Acknowledgements

This research received no external funding. This study only used publicly available GWAS summary statistics datasets. The author would like to thank Drs. Loscalzo and Cheng for insightful discussions and comments.

## Supplementary materials

**Figure S1.**
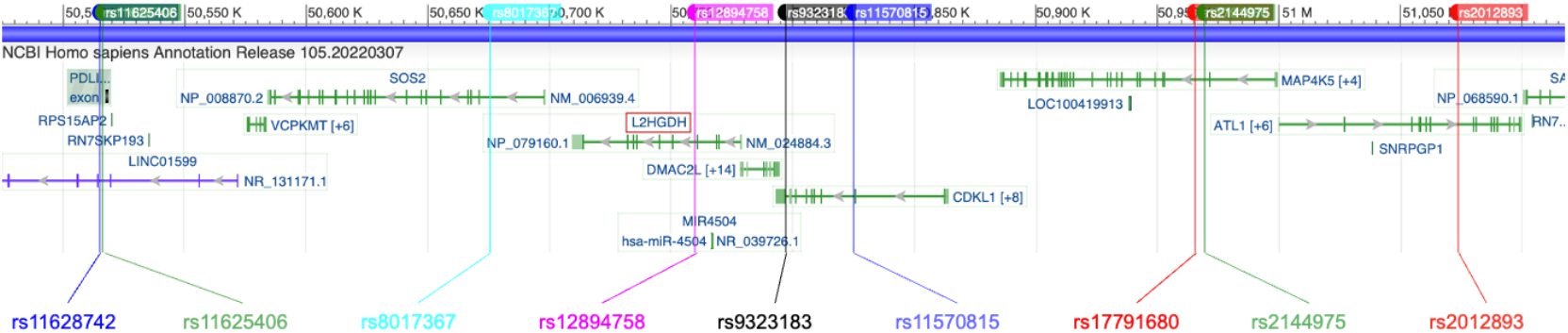
Genomic positions of nine genetic instruments for genetically proxied inhibition of *L2HGDH*. The genomic positions were visualized using the NCBI Graphical Sequence Viewer (GRCh37, accessed on Oct 29, 2022).

**Figure S2.**
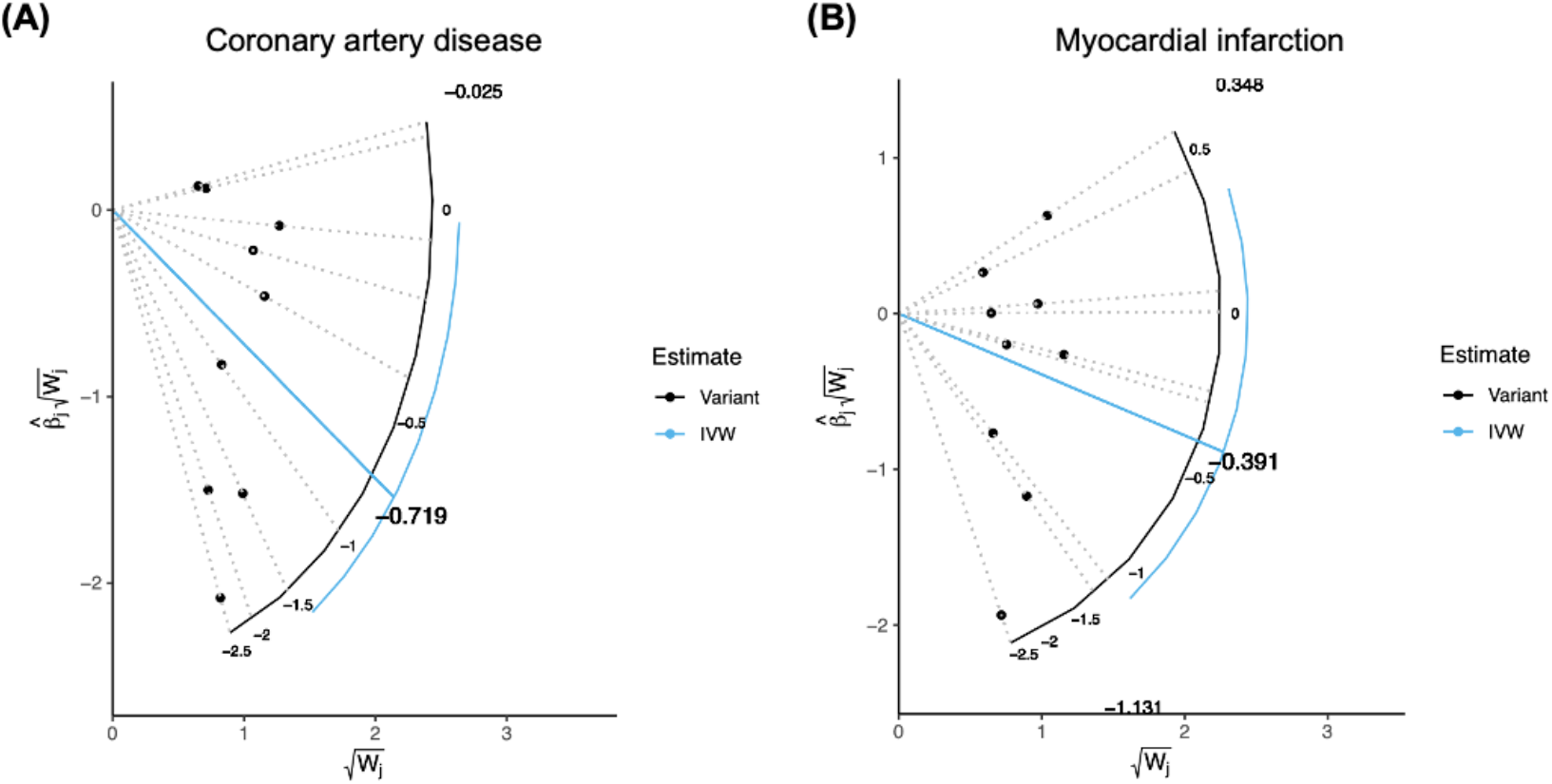
Radial inverse-variance weighted (IVW) Mendelian randomization plots for coronary artery disease (A) and myocardial infarction (B). Black dots indicate valid genetic variants. The estimated effect sizes are presented. There is no outlier variant in the present radial plots.

**Table S1.** Nine *L2HGDH*-proxied (± 500 kb) genetic variants that are associated with blood 2-hydroxyglutarate levels (P<0.05, r^2^<0.3).

| SNP | Position<br>(GRCh37) | EA/OA | EAF | $\beta$ (SE)* | P-value | F-statistic |
| --- | --- | --- | --- | --- | --- | --- |
| rs11628742 | chr14:50515075 | C/T | 0.438 | -0.0112 (0.0043) | $9.02 \times 10^{-3}$ | 6.784 |
| rs11625406 | chr14:50516192 | A/C | 0.619 | 0.0070 (0.0033) | $3.56 \times 10^{-2}$ | 4.500 |
| rs8017367 | chr14:50675487 | T/C | 0.261 | -0.0121 (0.0034) | $4.00 \times 10^{-4}$ | 12.665 |
| rs12894758 | chr14:50760030 | T/C | 0.291 | 0.0135 (0.0034) | $6.80 \times 10^{-5}$ | 15.766 |
| rs9323183 | chr14:50796881 | C/G | 0.054 | -0.0150 (0.0071) | $3.37 \times 10^{-2}$ | 4.463 |
| rs11570815 | chr14:50825258 | T/A | 0.662 | 0.0094 (0.0033) | $4.88 \times 10^{-3}$ | 8.114 |
| rs17791680 | chr14:50965445 | A/G | 0.372 | -0.0081 (0.0034) | $1.64 \times 10^{-2}$ | 5.676 |
| rs2144975 | chr14:50969448 | C/T | 0.955 | 0.0170 (0.0075) | $2.44 \times 10^{-2}$ | 5.138 |
| rs2012893 | chr14:51073443 | G/A | 0.088 | -0.0145 (0.0060) | $1.53 \times 10^{-2}$ | 5.840 |

SNP, single nucleotide polymorphism; EA, effect allele; OA, other allele; EAF, effective allele frequency.
\*Log(odds ratio) per 1-SD increase in blood 2-hydroxyglutarate levels.

**Table S2.** Genetic variants associated with blood 2-hydroxyglutarate levels (P<1×10^−5^, r^2^<0.3).

| SNP | Position<br>(GRCh37) | EA/OA | EAF | $\beta$ (SE)* | P-value |
| --- | --- | --- | --- | --- | --- |
| rs10196007 | chr2:145329033 | C/T | 0.184 | 0.0320 (0.0065) | $7.63 \times 10^{-7}$ |
| rs1568101 | chr3:148590090 | T/C | 0.912 | -0.0255 (0.0057) | $8.62 \times 10^{-6}$ |
| rs7725624 | chr5:63010725 | A/C | 0.087 | -0.0440 (0.0094) | $3.02 \times 10^{-6}$ |
| rs7702827 | chr5:158879869 | G/C | 0.700 | 0.0152 (0.0034) | $6.29 \times 10^{-6}$ |
| rs933341 | chr7:83743619 | G/A | 0.760 | -0.0166 (0.0035) | $1.56 \times 10^{-6}$ |
| rs8176023 | chr7:142642596 | G/A | 0.072 | 0.0411 (0.0087) | $2.46 \times 10^{-6}$ |
| rs890519 | chr8:18905049 | A/C | 0.416 | 0.0147 (0.0033) | $9.06 \times 10^{-6}$ |
| rs10966201 | chr9:24079188 | C/T | 0.345 | -0.0151 (0.0034) | $7.11 \times 10^{-6}$ |
| rs11793331 | chr9:133298199 | T/C | 0.024 | 0.0619 (0.0136) | $5.09 \times 10^{-6}$ |
| rs6484669 | chr11:11120287 | C/T | 0.259 | 0.0196 (0.0044) | $7.75 \times 10^{-6}$ |
| rs9544785 | chr13:78971070 | A/C | 0.361 | -0.0150 (0.0033) | $6.63 \times 10^{-6}$ |
| rs16977497 | chr17:71021645 | A/G | 0.045 | 0.0538 (0.0122) | $9.91 \times 10^{-6}$ |
| rs6064820 | chr20:58206004 | T/C | 0.723 | 0.0155 (0.0034) | $4.01 \times 10^{-6}$ |

SNP, single nucleotide polymorphism; EA, effect allele; OA, other allele; EAF, effective allele frequency.
\*Log(odds ratio) per 1-SD increase in blood 2-hydroxyglutarate levels.

**Table S3.** Phenotypes associated with the *L2HGDH*-proxied genetic instruments based on PhenoScanner V2 (accessed on Feb 6, 2023).

| SNP | Associated phenotypes ($P < 1 \times 10^{-5}$) |
| --- | --- |
| rs11628742 | - |
| rs11625406 | - |
| rs8017367 | - |
| rs12894758 | Impedance of arm right; Impedance of arm left |
| rs9323183 | - |
| rs11570815 | - |
| rs17791680 | Impedance of arm right; Impedance of arm left; Sitting height;<br>Arm fat-free mass right |
| rs2144975 | Treatment with inderal 10mg tablet |
| rs2012893 | - |

**Table S4.** Leave-one-out sensitivity analyses for the causal associations of genetically proxied *L2HGDH* inhibition with coronary artery disease and myocardial infarction using the inverse-variance weighted method.

| Removed SNP | Coronary artery disease |  | Myocardial infarction |  |
| --- | --- | --- | --- | --- |
|  | OR (95% CI) | P-value | OR (95% CI) | P-value |
| rs2144975 | 0.576 (0.277–1.194) | 0.138 | 0.826 (0.369–1.847) | 0.641 |
| rs11570815 | 0.545 (0.259–1.150) | 0.111 | 0.770 (0.337–1.758) | 0.535 |
| rs11625406 | 0.535 (0.260–1.103) | 0.090 | 0.715 (0.321–1.590) | 0.410 |
| rs17791680 | 0.499 (0.240–1.036) | 0.062 | 0.668 (0.297–1.499) | 0.328 |
| rs9323183 | 0.461 (0.225–0.945) | 0.035 | 0.644 (0.291–1.425) | 0.278 |
| rs2012893 | 0.457 (0.222–0.941) | 0.034 | 0.657 (0.296–1.461) | 0.304 |
| rs8017367 | 0.455 (0.212–0.979) | 0.044 | 0.553 (0.237–1.289) | 0.170 |
| rs11628742 | 0.445 (0.209–0.947) | 0.036 | 0.625 (0.271–1.442) | 0.270 |
| rs12894758 | 0.411 (0.188–0.899) | 0.026 | 0.648 (0.272–1.542) | 0.326 |
| None (original) | 0.486 (0.242–0.977) | 0.043 | 0.676 (0.312–1.463) | 0.320 |

SNP, single nucleotide polymorphism; OR, odds ratio; CI, confidence interval.

